# Purchase of hormonal contraceptive methods after delivery: a population-based study from Finland

**DOI:** 10.1101/2025.10.10.25337679

**Authors:** Camilla Jäntti, Elena Toffol, Jari Haukka, Oskari Heikinheimo

## Abstract

**Background:** Postpartum contraception start and use are important to avoid unwanted pregnancy and to optimize birth spacing. There is limited knowledge concerning the factors influencing the type and timing of contraceptive initiation and adherence after delivery.

**Methods:** This was a prospective register-based study. We identified 26 650 women that delivered between 1^st^ of January 2019 and 31^st^ of December 2019, using the Finnish Medical Birth Register. Postpartum follow-up time for each woman was one year. The primary outcome of the study was postpartum purchase of HC from a pharmacy, identified from the Prescription Centre. Additionally, we analyzed if mode of delivery, epidural analgesia, induction of labor, hypertensive complications, fear of childbirth or gestational diabetes affected HC purchase.

**Results:** Altogether 33% (n=8902) of the women purchased a HC method postpartum. The most purchased method was desogestrel-only pill (56%, n=4978), and other progestin-only preparations, which covered almost 90% of all methods started. Additionally, 4% (n=1141) received an intrauterine device (IUD) in primary health care. Instrumental delivery (OR 1.15, CI 1.06–1.25), cesarean section (OR 1.26, CI 1.13–1.41 for planned C-section and OR 1.25, CI 1.15–1.36 for emergency C-section), epidural analgesia (OR 1.12, CI 1.05–1.18) and induction of labor with balloon (OR 1.14, CI 1.01–1.28) were associated with a significantly higher rate of HC purchase.

**Conclusions:** Overall, 37% of women purchased a HC method or received an IUD during the first postpartum year. Common delivery complications were associated with higher rate of postpartum HC purchase.

## Introduction

Multiple international guidelines recommend a 12–24 months interpregnancy interval after childbirth. ^1–3^ Short interpregnancy interval has been associated with an increased risk of several adverse events, such as low birth weight, preterm delivery and maternal morbidity. ^4,5^ However, previous studies show that 7–39% of women have a short interpregnancy interval (defined here as another pregnancy within 18 months). ^4,6,7^ Altogether 55–70% of pregnancies after a short interpregnancy interval have been estimated to be unintended. ^6,8^ A recent systematic review found that more than half of women resume sexual intercourse within a couple of months after delivery. ^9^ Thus, accessing family planning services and the full range of contraceptive options and provision is crucial for enabling women to plan future pregnancies and optimize birth spacing.

Recommendations concerning the choice and timing of postpartum contraceptive methods, especially concerning combined hormonal contraception (CHC), vary considerably. According to the U.S guidance, the advantages of combined methods outweigh their risks already three weeks after delivery in non-breastfeeding women and after four weeks in breastfeeding women, ^10^ whereas the UK guideline recommends three and six weeks, respectively. ^3^ The Finnish guideline was updated at the beginning of 2025, recommending CHC is avoided for three months irrespective of breastfeeding status due to the risk of venous thromboembolism. ^11^ The previous version of the guideline, from 2015, recommended the use of progestin-only methods up to six months when breastfeeding. ^12^

There is limited knowledge concerning the factors influencing the type and timing of contraceptive initiation after delivery. Previous studies have shown that women express greater interest in using more effective contraception after high-risk pregnancies. ^13,14^ Most of these studies have combined multiple maternal and fetal risk factors and pregnancy complications and then compared high-risk versus low-risk pregnancies. A study from the U.S showed that women with gestational diabetes were more likely to receive permanent and long-acting reversible contraception (LARC). ^15^ Similarly another study reported that women with diabetes (pregestational and gestational) were more likely to use the most effective contraceptive methods after pregnancy, but the overall prevalence of contraceptive use did not differ between women with and without diabetes. ^16^ Little is known regarding the possible delivery-related factors that affect postpartum contraceptive choices. However female surgical sterilization is more common in association with cesarean section. ^17^

Aim of this study was to examine the rates and types of hormonal contraceptive methods purchased after delivery, using high quality Finnish registry data. Additionally, we examined the effects of pregnancy- and delivery-related factors on postpartum hormonal contraceptive purchase.

## Material and Methods

### Patient involvement

There was no patient involvement in the design of this study.

### Population

This is a prospective register-based cohort study from Finland. The original population included 294 356 users of hormonal contraceptives in 2017, selected from the Prescription Centre served by Kanta Services (for details, see https://www.kanta.fi/en/professionals/secondary-use-of-kanta-data), and 294 356 non-users of hormonal contraception, matched by age and municipality of residence. The matched non-users were randomly selected from the Social Insurance Institution database. The population has been described in detail in our previous papers. ^18,19^

The resulting cohort of 588 712 women living in Finland in 2017 was used as a sampling frame for this cohort study. From this population we identified all women who delivered (i.e., delivery at ≥ 22+0 weeks of gestational age) between January 1^st^ 2019 and December 31^st^ 2019 (n=26 650) (Appendix A).

### Registers

We gathered information on deliveries and delivery-related factors from the Finnish Medical Birth Register, managed by the Finnish Institute for Health and Welfare. This Register includes information on maternal age, previous deliveries and previous induced abortions, mode of delivery, epidural analgesia, induction of labor, gestational age, hypertensive complications, number of fetuses, fear of childbirth and gestational diabetes since 1987. Additionally, we gathered information on subsequent deliveries during the follow-up time from the Medical Birth Register and on abortions from the Finnish national Register of Induced Abortions.

The Prescription Centre includes information on all prescribed and redeemed medications in Finland. In this study we used Anatomical Therapeutic Chemical (ATC) codes for the hormonal contraceptive methods. We identified altogether 16 different ATC codes as first postpartum hormonal contraceptive prescriptions in the Prescription Centre (Appendix B). In the analyses we examined the total hormonal contraceptive purchase and thereafter divided the hormonal contraceptives in two groups: combined hormonal contraception (including pills, vaginal ring and patch) and progestin-only contraception (including pills, levonorgestrel-releasing intrauterine device (IUD) and contraceptive implants). Emergency contraception was excluded from the analyses.

We examined the IUD insertions in primary healthcare using the International Classification of Primary care (ICPC) code W12 (insertion of IUD). This code includes both copper and hormonal IUDs. This code is used only in some municipalities for primary healthcare in Finland, thus these data are not fully comprehensive.

### Variables

We used the following variables in the analyses: age, previous deliveries and induced abortions, mode of delivery, epidural analgesia, induction of labor, gestational age, hypertensive complications, number of fetuses, fear of the childbirth and gestational diabetes.

History of delivery was defined as one or more deliveries at a gestational age of more than 22 weeks. Induced abortion includes both medical and surgical induced abortions. We divided mode of delivery into four categories: vaginal delivery (including breech presentation), operative vaginal delivery (including vacuum and forceps), planned cesarean section and combined urgent and emergency cesarean sections. Epidural analgesia includes spinal analgesia, epidural analgesia and combined (spinal + epidural) analgesia. Induction of labor includes the use of intracervical balloon and prostaglandin (misoprostol) as the first induction method.

We divided gestational age into three groups: full-term (37 weeks or more), 33–37 weeks and less than 33 weeks. Hypertensive complications include the International Classification of Diseases (ICD-10) codes O14.0 (moderate pre-eclampsia), O14.1 (severe pre-eclampsia), O14.9 (undefined pre-eclampsia) and O13 (gestational hypertension without significant proteinuria). The ICD-10 code O99.80 is used for the fear of childbirth. In Finland oral glucose tolerance test (OGTT) is recommended by the national guideline on gestational diabetes for all pregnant women with risk factors for diabetes. OGTT is performed either between gestational weeks 12–16 and 24–28, or only between weeks 24–28. All cases of gestational diabetes were combined in the analyses irrespective of the type of management (diet, metformin, insulin or combined metformin and insulin).

The primary outcome of the study was the first purchase of hormonal contraception after the delivery. Secondary outcomes were an IUD insertion (either hormonal or copper) in primary healthcare, a subsequent delivery and an induced abortion. Each woman was followed-up for one year after the index delivery, or until outcomes, whichever came first.

### Statistical analysis

We described data using a cumulative incidence plot. We performed first univariable and multivariable logistic regression models with hormonal contraception purchase as the outcome of interest. Finally, we examined multinominal regression models to examine separately combined hormonal contraception and progestin-only contraception as an outcome. We performed all the statistical analyses using R software version 4.0.5. ^20^

## Results

Altogether 26 650 women of the study cohort delivered in Finland between January 1^st^ 2019 and December 31^st^ 2019. Women who purchased hormonal contraception after delivery were more likely primiparous, had an operative vaginal delivery or cesarean section as a delivery mode, had an epidural analgesia, hypertensive complications and fear of childbirth (Table 1).

**Table 1.**
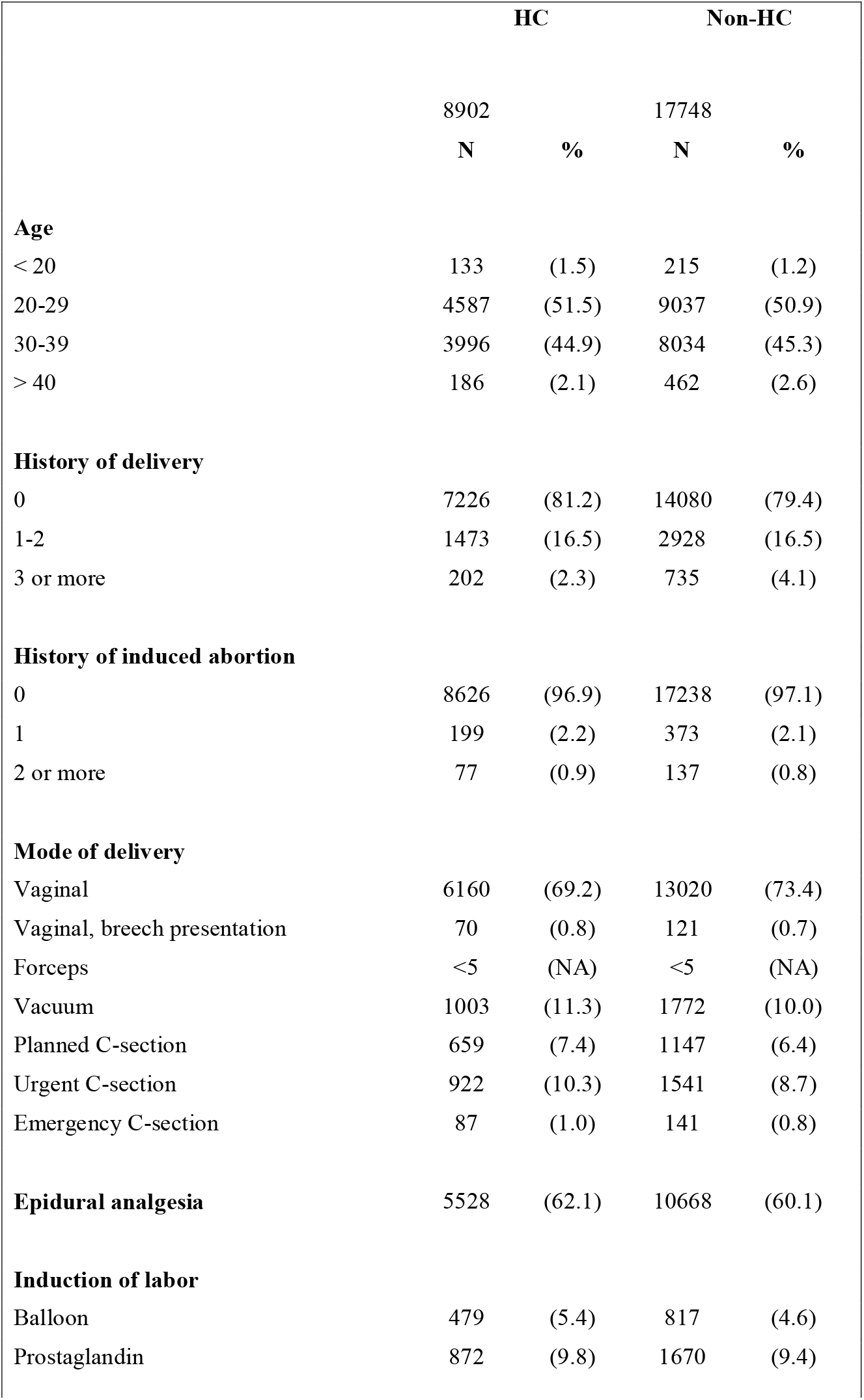
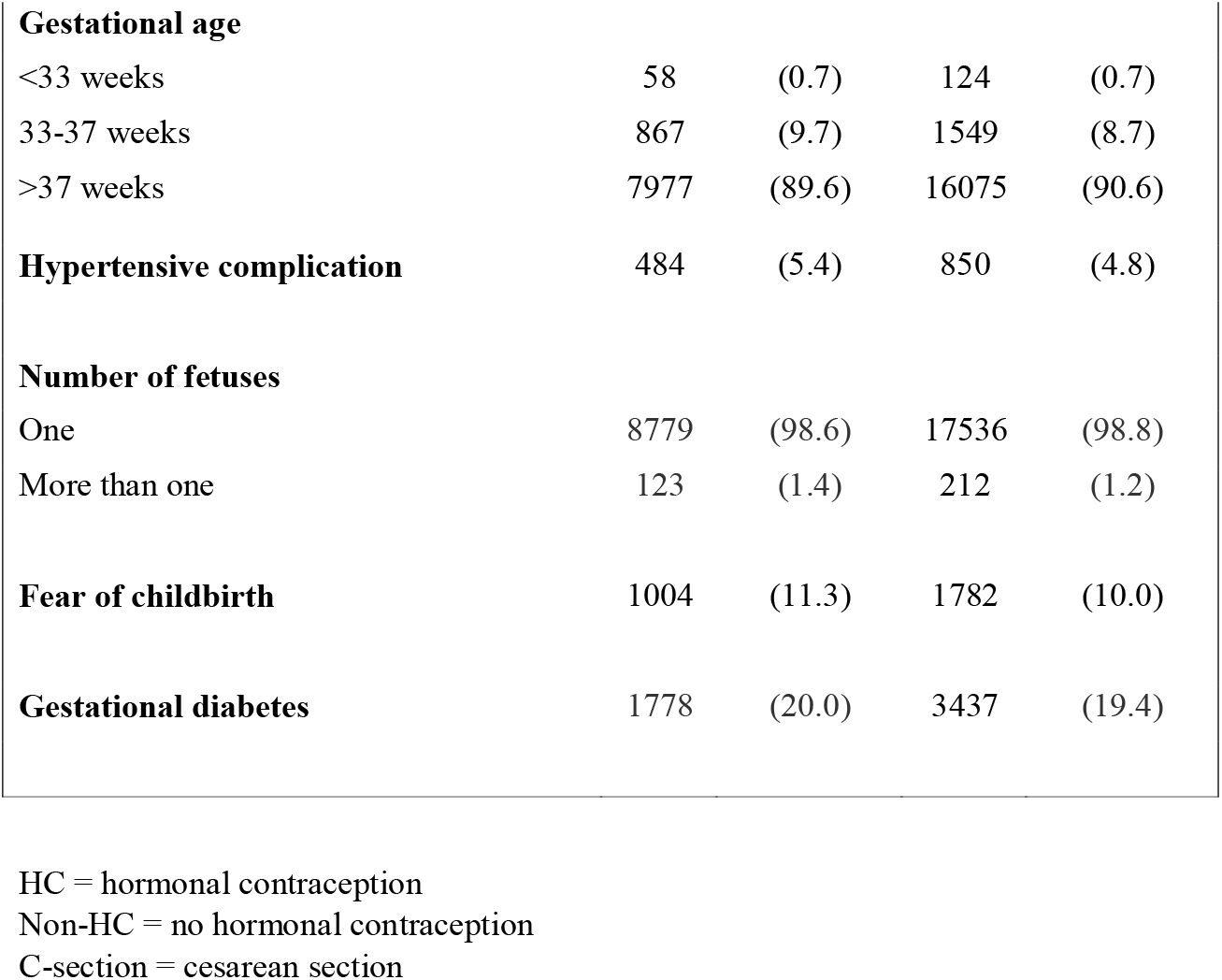
Basic characteristics of the study population according to purchased hormonal contraception after delivery in Finland, on year 2019.

According to the Prescription Centre 33% (n=8902) of the women purchased a hormonal contraceptive method within one year after the index delivery. The most often purchased method was desogestrel-only pill (56%, n=4978), followed by LNG-IUD (22%, n=1985) (Appendix B). Additionally, 4% (n=1141) received an IUD (either hormonal or copper) in primary health care. Of the study cohort 0.8% (n=204) underwent an induced abortion and 0.3% (n=76) delivered during the one-year follow-up (Figure 1).

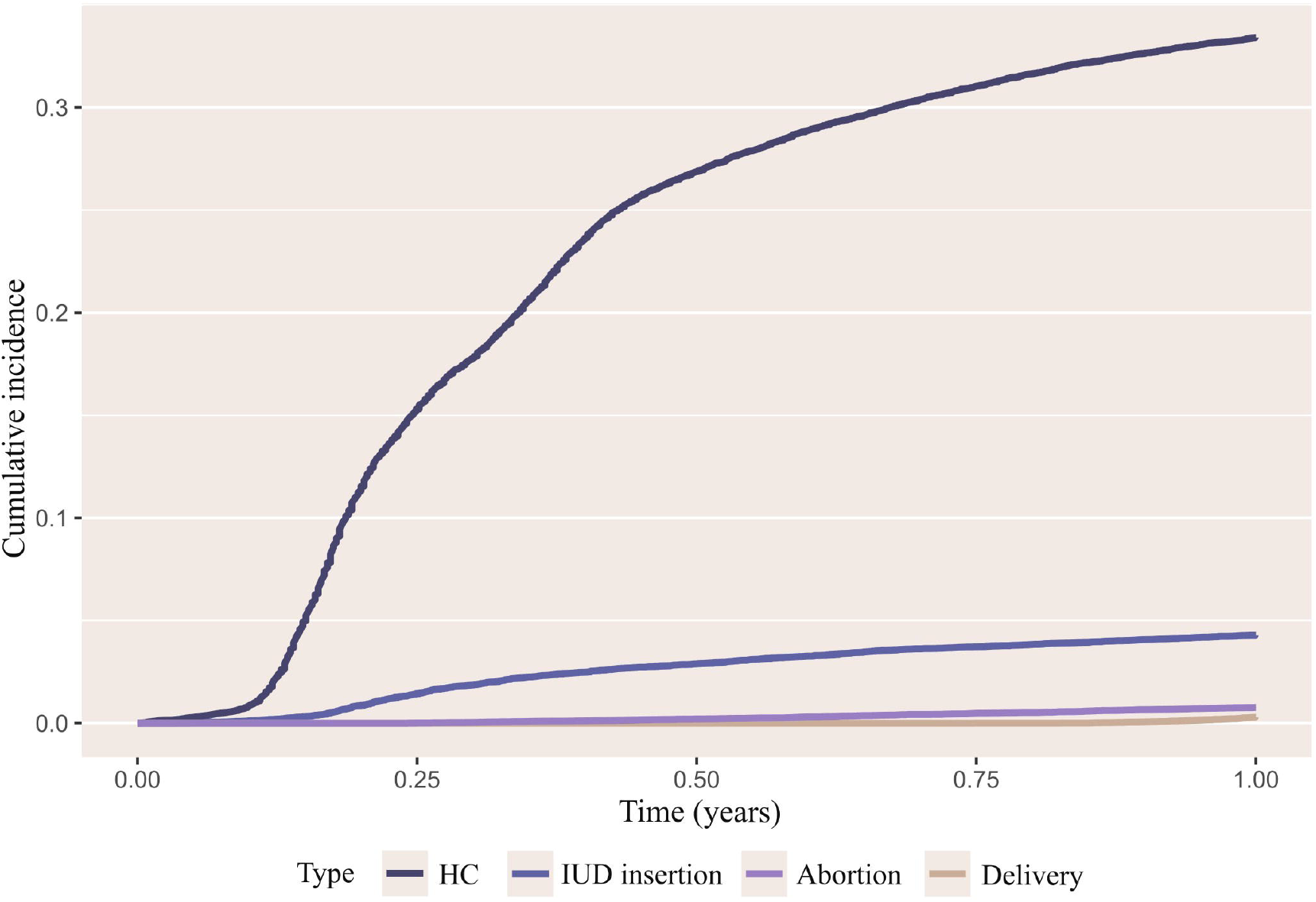

Age over 40 years and history of three or more previous deliveries were predictors of lower purchase of postpartum hormonal contraception (age > 40: odds ratio (OR) 0.70, 95% CI 0.53–0.93, previous deliveries > 3: OR 0.58, 95% CI 0.49–0.68). Higher odds ratio for purchasing hormonal contraception was associated with operative vaginal delivery (i.e. vacuum and forceps (OR 1.15, 95% CI 1.06–1.25), planned cesarean section (OR 1.26, 95% CI 1.13–1.41) and emergency cesarean section (OR 1.25, 95% CI 1.15–1.36)), use of epidural analgesia (OR 1.12, 95% CI 1.05–1.18) and induction of labor with balloon (OR 1.14, 95% CI 1.01–1.28) (Table 2).

**Table 2.** Odds ratios based on multinominal regression model for purchasing hormonal contraception after delivery according to reproductive history, pregnancy- and delivery-related factors in Finland, in the year 2019. Odds ratios (ORs) and 95% confidence intervals (CIs) for starting hormonal contraception after delivery overall (any HC) and divided into two groups (CHC and progestin-only). Information on purchased hormonal contraception was gathered from the Prescription Centre and on background factors from the Medical Birth Register.

|  | HC | CHC | Progestin-only |
| --- | --- | --- | --- |
| **History of delivery** |  |  |  |
| 0 | reference | reference | reference |
| 1-2 | 1.03 (0.96-1.11) | 0.95 (0.79-1.14) | 1.04 (0.97-1.12) |
| 3 or more | 0.58 (0.49-0.68) | 0.55 (0.34-0.87) | 0.58 (0.49-0.69) |
| **History of induced abortion** |  |  |  |
| 0 | reference | reference | reference |
| 1 | 1.07 (0.90-1.28) | 1.98 (1.43-2.73) | 0.95 (0.79-1.15) |
| 2 or more | 1.13 (0.85-1.49) | 2.16 (1.29-3.61) | 0.99 (0.73-1.35) |
| **Mode of delivery** |  |  |  |
| Vaginal | reference | reference | reference |
| Vacuum and forceps | 1.15 (1.06-1.25) | 1.38 (1.14-1.67) | 1.12 (1.02-1.22) |
| Planned C-section | 1.26 (1.13-1.41) | 2.27 (1.79-2.88) | 1.15 (1.02-1.30) |
| Emergency C-section | 1.25 (1.15-1.36) | 1.61 (1.32-1.95) | 1.21 (1.10-1.32) |
| **Epidural analgesia** | 1.12 (1.05-1.18) | 1.41 (1.21-1.63) | 1.09 (1.02-1.15) |
| **Induction of labor** |  |  |  |
| Balloon | 1.14 (1.01-1.28) | 0.95 (0.70-1.28) | 1.17 (1.03-1.32) |
| Prostaglandin | 1.03 (0.95-1.13) | 1.09 (0.88-1.35) | 1.03 (0.93-1.13) |
| **Gestational age** |  |  |  |
| < 33 weeks | reference | reference | reference |
| 33-37 weeks | 1.16 (0.84-1.61) | 0.70 (0.38-1.28) | 1.30 (0.91-1.85) |
| > 37 weeks | 1.04 (0.76-1.44) | 0.51 (0.28-0.91) | 1.20 (0.85-1.70) |
| **Hypertensive complication** | 1.07 (0.95-1.21) | 0.89 (0.67-1.19) | 1.10 (0.97-1.24) |
| **Age** |  |  |  |
| < 20 | reference | reference | reference |
| 20-29 | 0.83 (0.67-1.04) | 0.54 (0.37-0.79) | 0.92 (0.73-1.17) |
| 30-39 | 0.82 (0.66-1.03) | 0.36 (0.24-0.53) | 0.96 (0.76-1.23) |
| > 40 | 0.70 (0.53-0.93) | 0.18 (0.08-0.36) | 0.86 (0.64-1.16) |
| **Number of fetuses** |  |  |  |
| 1 | reference | reference | reference |
| 2 | 1.05 (0.83-1.32) | *0.90 (0.54-1.52) | *1.05 (0.82-1.34) |
| 3 | 0.34 (0.04-2.89) |  |  |
| **Fear of childbirth** | 1.08 (1.00-1.18) | 1.37 (1.15-1.65) | 1.04 (0.95-1.14) |
| **Gestational diabetes** | 1.03 (0.96-1.10) | 0.88 (0.75-1.03) | 1.05 (0.98-1.12) |

HC = hormonal contraception
CHC = combined hormonal contraception (combined oral contraceptives, vaginal ring, patch)
Progestin-only (progestin-only-pills, contraceptive implant, hormonal intrauterine device).
\* Number of fetuses is two or more

When analyzing the purchase of combined hormonal contraception and progestin-only contraception separately, the results were mostly similar to that of overall hormonal contraception. The main exception was higher odds ratio for purchasing combined hormonal methods if a woman had a history of induced abortion (one previous abortion (OR 1.98, 95% CI 1.43–2.73) and two or more previous abortions (OR 2.16, 95% CI 1.29–3.61)). Fear of childbirth was also associated with higher odds ratio for purchasing combined hormonal methods after delivery (OR 1.37, 95% CI 1.15–1.65). Odds ratio for purchasing combined hormonal methods was lower in all age groups above 20 years-of-age (i.e. 20–29 years (OR 0.54, 95% CI 0.37–0.79), 30–39 years (OR 0.36, 95% CI 0.24–0.53) and over 40 years (OR 0.18, 95% CI 0.09–0.36)) (Table 2).

## Discussion

We found that one third of the women purchased a hormonal contraceptive method in the year after delivery. More than half of them purchased desogestrel-only pills, and progestin-only preparations covered almost 90% of all the purchased methods. We also identified pregnancy- and delivery-related factors affecting the postpartum hormonal contraceptive purchase.

Previous studies have shown different rates of contraceptive use after delivery, varying from 46% to 83%. ^6,16,21,22^ Though, most of these studies include also non-hormonal and less effective methods. We were unable to include these methods to our register-based study. The rates of hormonal contraception initiation have been reported to be approximately 40–56%. ^6,22^ In the present study progestin-only preparations covered almost 90% of all the purchased methods. This result might be partly explained by Finnish guidelines, which recommend avoiding combined hormonal methods for a relatively long time after delivery. ^11^

In the present study the follow-up ended in an induced abortion for less than 1% of the cases. A Swedish study from 2020 showed an abortion rate of 2.5% within one to two years ^23^ and a study from China a 10% rate of abortion within two years after delivery. ^24^ Our results are likely to be lower given the follow-up ended when whichever outcome (i.e. contraceptive purchase or another pregnancy) was reached. In Finland almost 10% of all women undergoing an induced abortion have delivered during last two years. ^25^ Moreover, the overall incidence of induced abortion in Finland is low, approximately seven abortions per 1000 women of childbearing age (aged 15–49). ^25^

An operative vaginal delivery and a cesarean section were associated with an increased rate of purchasing hormonal contraceptive methods after delivery. This result was confirmed when examining combined hormonal contraceptives and progestin-only preparations separately. Previous studies have shown that women delay resuming sexual activity after assisted vaginal delivery or cesarean section. ^26^ It may be speculated that recovery takes longer after operative delivery, or women with instrumental delivery may wish to postpone another pregnancy with a reliable contraception. Some guidelines also recommend at least 12-month interpregnancy interval after a cesarean section, ^27^ even if the evidence of major complications such as uterine rupture in the next pregnancy is controversial. ^28,29^

Previous studies have reported high rates (78–82%) of postpartum contraceptive use in women with pregnancies complicated by diabetes. In these studies, women with diabetes preferred female surgical sterilization as a postpartum contraceptive method. ^15,30^ One found no difference in prevalence of postpartum contraceptive use between women with and without diabetes. ^15^ Similarly, in the present study gestational diabetes or hypertensive complications did not affect the rate of purchasing hormonal contraception after delivery.

We found that women older than 20 years-of-age purchased combined hormonal methods less than women younger than 20 years-of-age. This might be explained by Finnish guidelines, which recommend avoiding combined hormonal methods at least three months after delivery. ^11^ In this study we examined only the first prescription redeemed from a pharmacy. These women might change the contraceptive method later after delivery depending on their breastfeeding status and other medical conditions.

A major strength of the present nationwide study is that the cohort included more than half (51.7%) of women of fertile age living in Finland in 2017. ^31^ There were approximately 46 000 deliveries in Finland during 2019. ^32^ Thus, our study includes over half of all deliveries that year.

The study has also some limitations. Many women might rely on lactation amenorrhea as a contraceptive method or start with a condom as their first choice after delivery. We were unable to include information on breastfeeding status, as this information is not recorded to the Medical Birth register. Breastfeeding has an effect on the choice of the method, as progestin-only methods are recommended by the Finnish guidelines for three months when breastfeeding. ^11^ Our study also failed to include non-hormonal methods such as condoms, most copper-IUDs and sterilizations.

In addition, our results on slightly lower uptake of hormonal contraceptive methods might be partly explained by missing data due to the contraceptive methods provided free-of-charge. Many Finnish municipalities offer free-of-charge contraception to young women (usually under 20 or 25 years-of-age). At the time of the study some municipalities also offered first LARC method or LARC after delivery, regardless of age. ^33^ Because information on free-of-charge hormonal contraception postpartum is not fully recorded in the Prescription Centre, we were unable to include this information. This limitation was partly mitigated by including data on insertion of intra-uterine devices in primary healthcare using the relevant ICPC code.

Moreover, we had no information on whether the women in fact started the purchased hormonal contraceptive method. In this study the primary outcome was a redeemed hormonal contraceptive prescription based on the Prescription Centre data. However, as hormonal contraception is not reimbursed in Finland, we assumed that most of the women who redeemed their prescription did start using hormonal contraceptive method when redeemed.

In conclusion, one third of the women purchased hormonal contraception in the year following the delivery. Moreover, several pregnancy- and delivery-related factors affecting the contraceptive purchase after delivery can be recognized. However, majority did not start any hormonal contraception and as mentioned before 10% of Finnish women undergoing an induced abortion have delivered during the last two years. There is an unmet need for contraception in this group of women. Information on contraceptive initiation and background factors affecting it helps to tailor contraceptive counselling even more towards an individual and women-centered approach. Future research is needed to examine the compliance of started contraception, and factors affecting it during long-term follow-up.

## Supporting information

Appendix A, Flowchart

Appendix B, ATC codes

## Data Availability

All data produced in the present study are available upon reasonable request to the authors.

## Acknowledgements

We thank Jennifer Rowland for language revision of this article.

## Funding

This work was supported by the Jane and Aatos Erkko Foundation (JH, grant number 170062), the Avohoidon tutkimussäätiö (JH, Foundation for Primary Care Research), the Yrjö Jahnsson Foundation (ET, grant number 20207328), Finnish Cultural Foundation (ET, grant number 00211101 and 00230159) and Helsinki University Central Hospital HUS (CJ).

## Author contributions

**Camilla Jäntti:** Conceptualization, Formal analysis, Writing – Original Draft. **Elena Toffol:** Conceptualization, Formal analysis, Writing - Review & Editing, Supervision. **Jari Haukka:** Conceptualization, Methodology, Software, Formal analysis, Writing - Review & Editing, Supervision. **Oskari Heikinheimo:** Conceptualization, Writing - Review & Editing, Supervision.

**Appendix A: Flowchart of the study population**

**Appendix B: Supplementary table of ATC codes**

## Notes

### Competing Interest Statement

CJ, ET and JH none
OH serves occasionally on scientific advisory boards organized by Bayer AG and Gedeon-Richter, and has lectured at educational events organized by these companies.

### Funding Statement

This work was supported by the Jane and Aatos Erkko Foundation (JH, grant number 170062), the Avohoidon tutkimussaatio (JH, Foundation for Primary Care Research), the Yrjo Jahnsson Foundation (ET, grant number 20207328), Finnish Cultural Foundation (ET, grant number 00211101 and 00230159) and Helsinki University Central Hospital HUS (CJ).

### Author Declarations

Ethics Committee of the Faculty of Medicine, University of Helsinki reviewed this study (3/2018). Because this is a register-based study, no individual consent is needed.

