## Appendix A, Flowchart for "Purchase of hormonal contraceptive methods after delivery: a population-based study from Finland"

### Slide 1
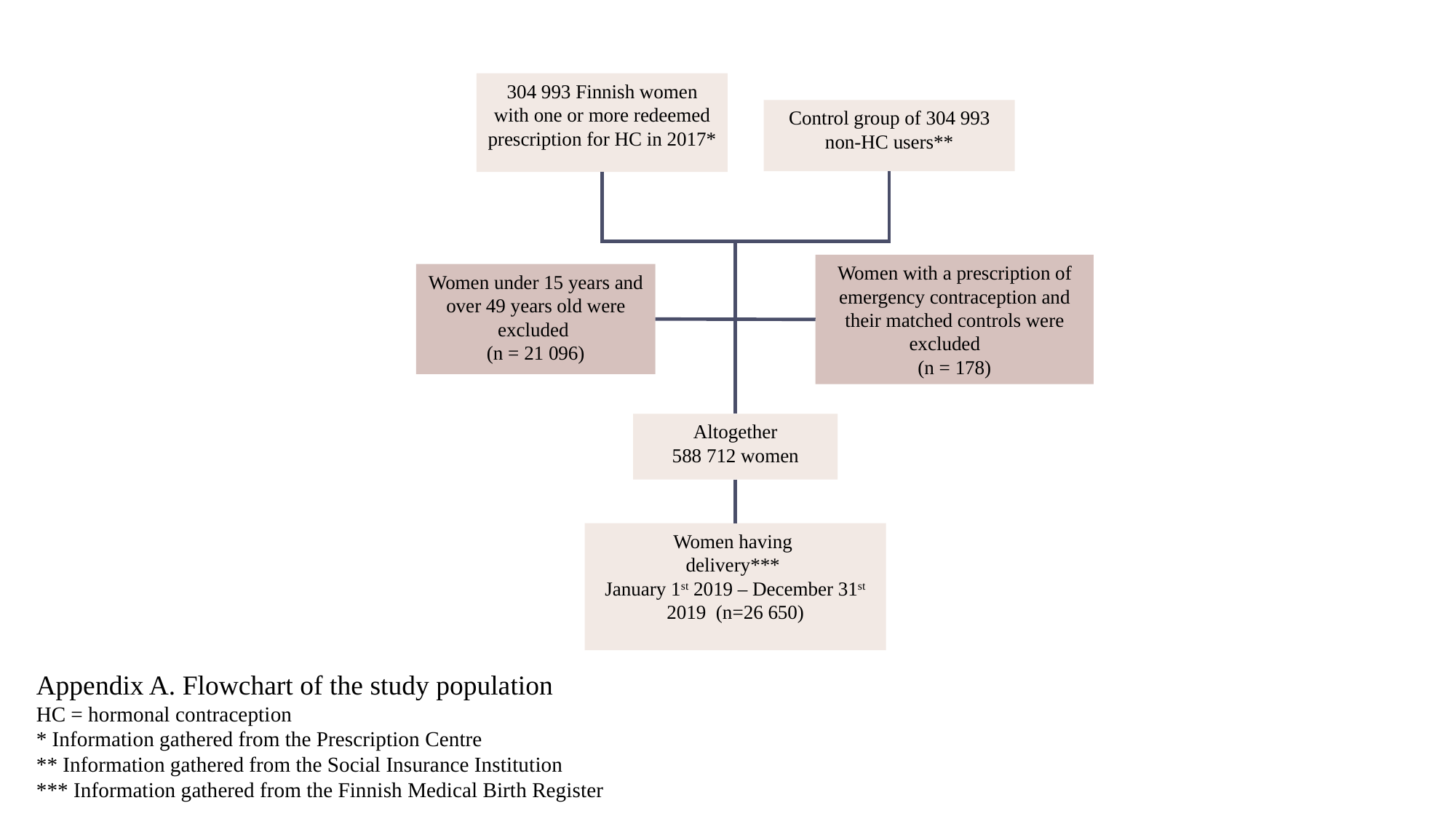

304 993 Finnish women with one or more redeemed prescription for HC in 2017*
Control group of 304 993 non-HC users**
Women with a prescription of emergency contraception and their matched controls were excluded
(n = 178)
Women under 15 years and over 49 years old were excluded
(n = 21 096)
Altogether
588 712 women
Women having
delivery***
January 1^st^ 2019 – December 31^st^ 2019 (n=26 650)
Appendix A. Flowchart of the study population
HC = hormonal contraception
* Information gathered from the Prescription Centre
** Information gathered from the Social Insurance Institution
*** Information gathered from the Finnish Medical Birth Register
