## Appendix B, ATC codes for "Purchase of hormonal contraceptive methods after delivery: a population-based study from Finland"

**Appendix B. ATC codes and preparations of hormonal contraceptive methods started after a delivery in Finland, 2019-2020, according to the Prescription Centre.**

| **ATC code and preparation** | **8902** | **%** |
| --- | --- | --- |
| **Combined oral contraception** |  |  |
| **Fixed combinations** |  |  |
| G03AA07 = levonorgestrel + EE | 12 | 0.13 |
| G03AA09 = desogestrel + EE | 102 | 1.15 |
| G03AA10 = gestodene + EE | 144 | 1.62 |
| G03AA12 = drospirenone + EE | 415 | 4.66 |
| G03AA14 = nomegestrol + estradiol | 46 | 0.52 |
| G03AA16 = dienogest + EE | 35 | 0.39 |
| G03HB01 = cyproterone + EE | 73 | 0.82 |
| **Sequential preparations** |  |  |
| G03AB08 = dienogest + estradiol | 42 | 0.47 |
| **Vaginal ring** |  |  |
| G02BB01 = etonogestrel + EE | 165 | 1.85 |
| **Patch** |  |  |
| G03AA13 = norelgestromin + EE | 83 | 0.93 |
| **Progestin-only-pills** |  |  |
| G03AC01 = norethisterone | 327 | 3.67 |
| G03AC09 = desogestrel | 4978 | 55.93 |
| G03AC10 = drospirenone | 45 | 0.51 |
| **Implant** |  |  |
| G03AC08 = etonogestrel | 325 | 3.65 |
| **LNG-IUD** |  |  |
| G02BA03 = levonorgestrel | 1985 | 22.30 |
| G03AC03 = levonorgestrel** | 124 | 1.39 |

8902 women redeemed at least one prescription of hormonal contraceptive method after delivery during a one-year follow-up.

ATC code = Anatomical Therapeutic Chemical code (https://www.whocc.no/atc_ddd_index/)

EE = ethinylestradiol

LNG-IUD = levonorgestrel-releasing intrauterine device

*ATC code G03HB01 (cyproterone + ethinylestradiol) was included in the combined hormonal contraception group, as this combination is commonly used in Finland.

**ATC code G03AC03 is levonorgestrel for systemic use. This ATC code includes one pill and one implant.
